# Identifying differences between those with suicidal ideation-with-action, compared to ideation alone, using a community representative sample

**DOI:** 10.1101/2024.12.25.24319634

**Authors:** C Chan, W Wodchis, P Kurdyak, P Donnelly

## Abstract

**Background:** Few studies examine suicidal ideation in the general population and who might act on suicidal thoughts. It is important to understand ideators, the largest group on the suicidality continuum.

**Objectives:** This study examines factors associated with suicidal ideation among community-dwelling individuals, and sociodemographic, health and help-seeking factors associated with ideation accompanied by planning or suicide attempt (‘ideation-with-action’) compared to ideation alone.

**Methods:** Using the 2002 and 2012 Canadian Community Health Surveys – Mental Health cycles (CCHS-MH), this cross-sectional cohort study examined 14,708 Ontarians 15 years and older who answered questions about suicidal ideation, and compared characteristics between non-ideators, ideators with a plan or previous attempt, and ideators alone, with chi-square tests and logistic regression.

**Results:** 2.1% of CCHS respondents reported past-year ideation alone (n=302) and another 0.5% reported ideation with plan or past-year suicide attempt (n=76).

The risk profile of ideators compared to non-ideators was similar to that of ideators-with-action compared to ideators-without-action: male, younger, unpartnered, less educated, have lower income, no job, have a mood and anxiety disorder, a substance use disorder and seek help for mental health problems.

Most ideators (65%) do not seek help, and those with a plan or previous suicide attempt are more likely to do so.

**Conclusion:** Ideators differ in profile in terms of whether they have ideation only, have made a plan or had previous attempts. Risk factors differentiating ideators from non-ideators are the same factors that further differentiate ideators-with-action compared to those with only ideation, suggesting the existence of a suicidality continuum and opening up the opportunity for targeting common risk factors in prevention efforts.

## Introduction

Suicidal ideation is a precursor to suicide and the most important initial stage of suicidality. It is the third most important predictor of suicide, behind mental health hospitalizations and suicide attempts (Franklin et al, 2017; World Health Organization, 2014). However, only a small proportion of those with ideation will act on their thoughts (Nock et al, 2008a; Maris, 1981). It is, therefore, a significant predictor but not a very discriminatory predictor, like most other suicide risk factors.

In an effort to understand ideation and its progression to action or not, contemporary suicidality models distinguish suicidal ideation from suicidal behaviour and use ‘ideation-to-action’ models to emphasize that risk factors for ideation differ from risk factors for suicide attempt (Klonsky et al, 2018). A systematic review found that the rate of transitioning to a suicide attempt among people with ideation ranges between 2.6% and 37% (Haregu et al, 2023). However, studies differentiating those with ideation from those who have attempted or will attempt are rare (May & Klonsky, 2016; Glenn and Nock, 2014), and where studies exist, they tend to focus on subpopulations such as veterans (Veterans Affairs Canada, 2019) or students (Liu et al, 2023) and do not provide general community-based estimates.

This study sought to advance understanding of community-dwelling persons with ideation and characteristics that distinguish those who only think about suicide from those who act on their suicidal thoughts in a general population sample. The study explored sociodemographic factors, health status factors including presence of mood/anxiety/substance use disorders, and help-seeking behaviour to ascertain which variables were more strongly associated with those who have planned or attempted suicide, compared to those who have ideation alone.

### Research objectives

The study had two objectives. First, to compare individuals with suicidal ideation to those with no suicidal ideation on a number of sociodemographic, health and help-seeking characteristics. Second, to compare individuals with suicidal ideation accompanied by a plan or a previous attempt (ideation-with-action) to those with suicidal ideation alone.

## Methods

### Study population

The study population was created by pooling Ontario respondents from two surveys: the 2002 and 2012 Canadian Community Health Surveys – Mental Health (CCHS-MH) administered by Statistics Canada between May and December 2002, and between January and December 2012. The overall survey response rate was 77% for the 2002 survey (CCHS, 2002) and 69% for the 2012 survey (CCHS, 2013). The study population was restricted to CCHS respondents who provided consent to linkage with health administrative data (79%).

### Data source

The CCHS-MH was a cross-sectional survey conducted in 2002 and 2012 by Statistics Canada that used a multistage sample allocation strategy to gather data from a randomly selected representative sample of households across Canada. It included modules on health status, mental well-being, mental illness, determinants of mental health, demographics and other variables (Statistics Canada, 2014; Beland, 2002). Participants were aged 15 years and older living in private dwellings in Canada’s ten provinces covering approximately 98% of Canada’s population. Detailed methodology is available from Statistics Canada reports (2004, 2013a, 2013b, 2014).

### Cohort creation

All Ontario CCHS-MH respondents from 2002 and 2012 survey cycles were considered, and after removing those without consent for linkage and those with missing data for suicidal ideation, 14,708 were included in the analysis (**Figure 3.1**).

**Figure 3.1:**
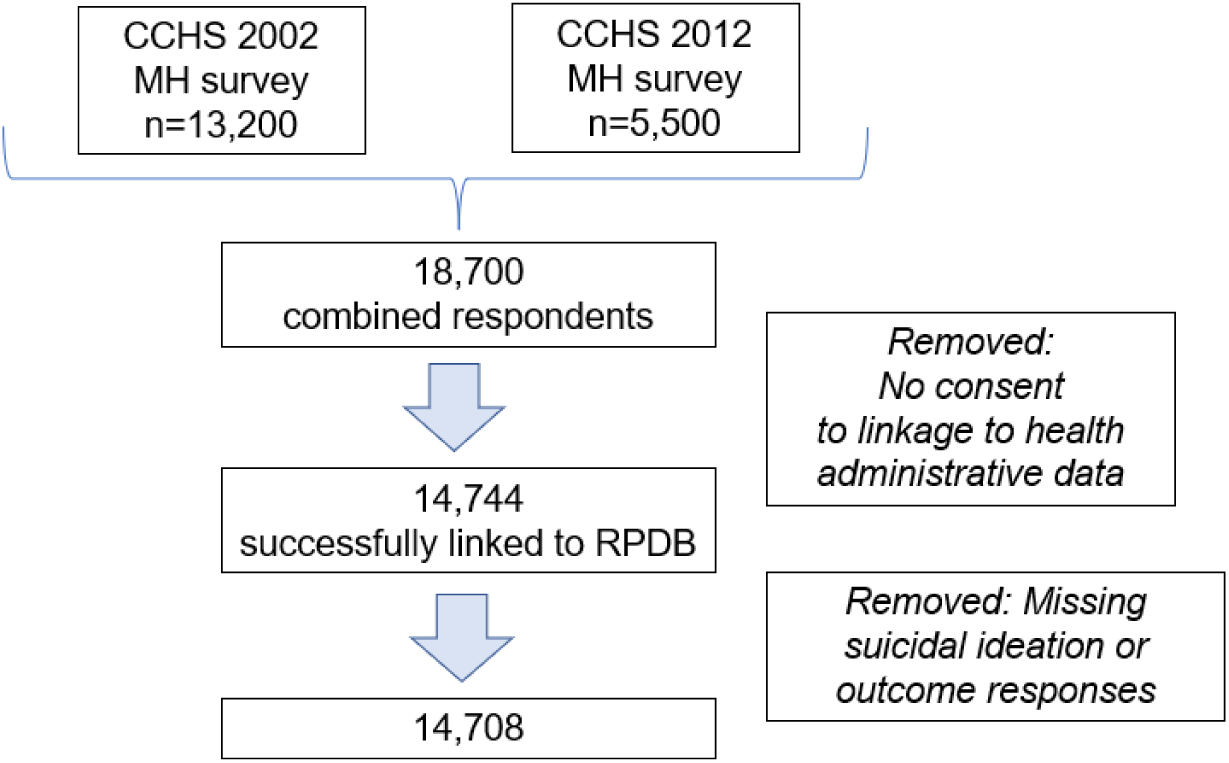
Flow chart of study population from pooled Ontario component of Statistics Canada’s Canadian Community Health Surveys (CCHS) Mental Health component

### Determination of subgroups

Subgroups were identified as follows.

*i. Suicidal ideation (past 12 months)*: All CCHS respondents were asked the question about lifetime suicidal ideation “Have you ever seriously thought about committing suicide or taking your own life?” and if they answered yes, then they were asked *“Has this happened in the past 12 months?”* with two response options: *Yes, No.* For this study, respondents were classified as reporting ideation if they answered “yes” to the latter question.

In the CCHS, respondents were asked about suicidal ideation only if they answered yes to a subclinical depression question “Did you feel sad, empty or depressed most of the day, nearly every day, during (a) period of 2 weeks?”.

i. *ii. Suicidal ideation with plan*: All CCHS respondents who answered affirmatively to question (i) were asked the question *“In the past 12 months”, “you made a plan for committing suicide?”* with two response options: *Yes, No.* Respondents were classified as having ideation with a plan if they responded “yes”.
ii. *Suicidal ideation with past-year suicide attempt:* All CCHS respondents who answered affirmatively to question (i) were asked the question *“During the past 12 months”, “you attempted suicide or tried to take your own life?”* with two response options: *Yes, No.* Respondents were classified as having ideation with a past-year attempt if they responded “yes”.

These responses were first used to create two subgroups in this study are:

1. Those without suicidal ideation; and
2. Those with suicidal ideation.

This latter group was further subdivided for analysis into those with ideation alone and those with ideation-with-action (individuals with a plan or past-year suicide attempt).

### Covariates

Covariates were the following baseline characteristics from the CCHS survey.

Sociodemographic covariates were: **age, sex, marital status, income, education and employment status**. These variables were chosen as they have been shown to be important risk factors for suicidal ideation and behaviour (for age: WHO, 2014; Patton et al, 2009; Nock et al, 2008a; for sex: Park et al, 2005; Ibrahim et al, 2017; Lewinsohn et al, 2001; for marital status: Kolves et al, 2010; Yeh et al, 2008; Brockington, 2001; for income: McMillan et al, 2010; Lynch et al, 2000; Ferrada-Noli, 1997; for education: Phillips and Hempstead, 2017; Lorant et al, 2005; for employment: Milner et al, 2014; Milner et al, 2013). Detailed evidence in the literature justifying the selection of covariates can be found in **Appendix 1**.

In this study, age was divided in the following categories: 15-24, 25-44,45-64, 65-84, 85 and above. Sex was male or female. Income was analyzed in the following categories and expressed in Canadian dollars: $0-$39,999, $40,000-$59,999, $60,000 and above. Education was analyzed in the following categories: less than secondary school, completed secondary school and post-secondary school. Employment was analyzed as having a job in the past year versus not. Marital status was classified as either “single, widowed, separated, divorced” (grouped together) versus “married or common-law” (grouped together).

Two health status covariates were analysed for their impact on any association between suicidal ideation and the study outcomes:

i. **Mood and anxiety disorders** were defined by using the Diagnostic and Statistical Manual of Mental Disorders, IV Edition (DSM-IV) criteria (American Psychiatric Association, 2000) and were assessed in the CCHS by using a Canadian adaptation of the World Mental Health Composite International Diagnostic Interview (WMH-CIDI), which is a structured interview for psychiatric disorders (Statistics Canada, 2013a; Kessler and Ustun, 2004). Diagnostic algorithms identified respondents meeting the criteria for the measured mood and anxiety disorders (major depressive episode, bipolar I, bipolar 2, hypomania, or phobia, obsessive-compulsive disorder, agoraphobia, panic disorder, social phobia) in the past 12 months. For example, a diagnosis of major depressive episode requires at least one episode of two weeks or more of persistent low mood, loss of interest or pleasure in normal activities with associated changes in sleep pattern, changes in appetite, feelings of guilt, hopelessness, impaired concentration, or suicidal thoughts (Statistics Canada, 2013b).
ii. **Substance use disorder** includes any alcohol and drug use disorder and was measured using the modified WMH-CIDI algorithms derived from DSM-IV applied to symptom data to define substance use disorders, that is, substance abuse and/or dependence for alcohol, cannabis and other drugs, in the past 12 months. Substances covered were cannabis, cocaine, amphetamines, MDMA (ecstasy), hallucinogens, solvents, heroin, and steroids.

Lastly, **self-reported help-seeking** was also studied as a covariate. The CCHS collected information about respondents’ use of help, and health care services related to mental health problems (i.e. problems with emotions, mental health, or use of alcohol or drugs) during the past 12 months through this question “Have you ever seen, or talked on the telephone, to any of the following professionals about your emotions, mental health or use of alcohol or drugs?” which was asked of all respondents. The professionals included psychiatrists, family doctors or general practitioners, other medical doctors, psychologists, nurses, social workers, counsellors or psychotherapists, religious or spiritual advisors, and other professionals. “Other professionals” referred to acupuncturists, biofeedback teachers, chiropractors, energy healing specialists, exercise or movement therapists, herbalists, homeopaths or naturopaths, hypnotists, guided imagery specialists, massage therapists, relaxation experts, yoga or meditation experts, and dieticians.

### Statistical analysis

Baseline characteristics of persons without ideation, those with ideation only, and those with ideation-with-action, were compared using unweighted frequencies and weighted percentages for categorical variables. Chi-square tests were used to compare difference in proportions between the groups since all covariates were categorical variables. Statistical significance was indicated by a two-tailed *P* value < 0.05.

For the first objective, a logistic regression model examined associations with suicidal ideation (reference: no suicidal ideation) and measured the strength of associations with the covariates. For the second objective, a logistic regression model examined associations with suicidal ideation-with-action compared to suicidal ideation alone and measured the strength of associations with the covariates. Covariates significantly associated with suicidal ideation in univariate analyses were included in these multivariate analyses.

Specifically, models were run comparing these groups:

1. Those with ideation versus those without ideation. Results are shown in **Tables 3.1 and 3.2**.
2. Those with ideation alone versus those with ideation-with-action (i.e. accompanied by either a plan or past-year attempt). Results are shown in **Tables 3.3** and **3.4**.

The possibility of multicollinearity between the variables of interest was examined using the variance inflation factor (VIF) indicator; there is no indication of multicollinearity as the VIF of all variables of interest was <5 (Kim, 2019). Correlation matrices were also examined, and elimination and combination of variables tested, to assess for collinearity. Hosmer-Lemeshow goodness-of-fit tests were conducted to test the fit of the regression models.

Sampling weights provided by Statistics Canada were applied in all analyses to account for the multistage survey sampling design and to ensure that estimates were representative of the Ontario population.

Sensitivity analyses to examine the impact of any differences in the two surveys, and the impact of excluding observations with missing data (see **Appendix 2**.)

All analyses were performed with SAS version 9.4 (SAS Institute Inc, Cary, North Carolina).

### Ethics

The survey datasets used in this study were accessed at Institute for Clinical Evaluative Sciences (ICES), an independent, non-profit research institute in Ontario which uses population-based health information to produce knowledge on a wide range of healthcare issues. The study cohort consists of CCHS respondents who consented to have their survey data shared with ICES for linkage with health administrative data. The use of these data for health system planning and evaluation was authorized under Section 45 of Ontario’s Personal Health Information Protection Act, which allows for administrative approval and does not require review by a research ethics board.

Data for the study was accessed for research purposes from 26 April 2023 to 26 September 2024. Authors did not have access to information that could identify individual participants during or after data collection.

## Results

Persons reporting ideation versus those with no ideation were examined. Baseline characteristics of these two groups are presented in **Table 3.1.**

**Table 3.1.** Baseline characteristics of those with suicidal ideation and the referent group (those without ideation) *.

|  |  | No suicidal ideation<br>N=14,330 (97.4%) |  | Suicidal ideation<br>N=378 (2.6%) |  | p |
| --- | --- | --- | --- | --- | --- | --- |
|  |  | n | weighted<br>% | n | weighted<br>% |  |
| **Age group** |  |  |  |  |  | **0.0001<sup>a</sup>** |
|  | 15-24 | 2202 | 15.9 | 106 | 26.6 |  |
|  | 25-44 | 4695 | 35.6 | 125 | 39.2 |  |
|  | 45-64 | 4202 | 32.2 | 110 | 27.6 |  |
|  | 65-84 | 2980 | 15.1 | 37 | 6.6 |  |
|  | 85 and above | 251 | 1.2 | 0 | 0.0 |  |
| **Sex** |  |  |  |  |  | **0.93** |
|  | Male | 6626 | 50.0 | 182 | 48.6 |  |
|  | Female | 7704 | 50.0 | 196 | 51.4 |  |
| **Income** |  |  |  |  |  | **0.0083** |
| | \$0-\$39,999 | 8869 | 53.0 | 254 | 63.6 | |
| | \$40,000-\$59,999 | 2329 | 17.6 | 45 | 11.4 | |
| | \$60,000 and over | 1714 | 11.6 | 33 | 5.7 | |
|  | Not stated | 1418 | 17.8 | 46 | 19.3 |  |
| **Past-year employment** |  |  |  |  |  | **0.41** |
|  | Employed | 9339 | 70.9 | 250 | 70.5 |  |
|  | Unemployed | 3664 | 22.9 | 114 | 26.8 |  |
| **Education** |  |  |  |  |  | **0.0904** |
|  | Less than secondary school | 3514 | 20.6 | 114 | 27.8 |  |
|  | Secondary school | 2619 | 17.7 | 68 | 17.4 |  |
|  | Post-secondary | 8137 | 61.7 | 196 | 54.8 |  |
| **Marital Status** |  |  |  |  |  | **<0.0001** |
|  | Married or Common Law | 6874 | 61.4 | 113 | 40.6 |  |
|  | Single/Widowed/Separated/Divorced | 6588 | 38.6 | 253 | 59.4 |  |
| **Substance Use** |  |  |  |  |  | **<0.0001** |
|  | yes | 434 | 3.1 | 58 | 16.0 |  |
|  | no | 13736 | 95.9 | 310 | 81.2 |  |
|  | Don't know / Not stated | 160 | 1.1 | 10 | 2.8 |  |
| **Mood and anxiety disorder** |  |  |  |  |  | **<0.0001** |
|  | yes | 1356 | 8.8 | 130 | 40.6 |  |
|  | no | 12728 | 90.3 | 238 | 58.4 |  |
|  | Don't know / Not stated | 246 | 0.9 | 10 | 1.0 |  |
| **Substance Use & mood and anxiety** |  |  |  |  |  | **<0.0001** |
|  | yes | 113 | 0.7 | 26 | 7.8 |  |
|  | no | 12282 | 87.1 | 202 | 48.6 |  |
| **Seek help** |  |  |  |  |  | **<0.0001** |
|  | yes | 67 | 0.5 | 77 | 34.7 |  |
|  | no | 14263 | 99.5 | 301 | 65.4 |  |
| *\* Notes:* |  |  |  |  |  |  |
| *Counts are unweighted.* |  |  |  |  |  |  |
| *Percentages and p values are weighted.* |  |  |  |  |  |  |
| <sup>a</sup> p-value calculated on 15 to 84 age groups |  |  |  |  |  |  |

2.6% of community-dwelling survey respondents (n=378) reported suicidal ideation in the past year.

As shown in **Table 3.1**, individuals with suicidal ideation tended to be younger, poorer, unpartnered, have a substance use disorder, mood and anxiety disorder, and have sought professional help (p < 0.01). There were no differences between those with and those without suicidal ideation in terms of sex, past-year employment and education.

Logistic regression results (**Table 3.2**) show all variables having statistically significant associations with ideation. The strongest factors associated with increased risk of suicidal ideation were help-seeking (aOR = 32.1, 95% CI: 20.9, 49.4), having a substance use disorder (aOR = 3.3; 95% CI: 3.2, 3.3), having a mood and anxiety disorder (aOR = 2.9; 95% CI: 2.8, 2.9), and being younger (aOR=2.9; 95% CI 2.9, 3.0). Marriage or common-law relationships was a protective factor against ideation (aOR = 0.5; 95% CI: 0.5, 0.5).

**Table 3.2.** Association of sociodemographic factors, health conditions, and help-seeking

|  |  | No ideation (n=14,330) vs Suicidal ideation (n=378) |  |
| --- | --- | --- | --- |
|  |  | Crude | Adjusted |
|  |  | OR (95% CI) | OR (95% CI) |
| **Sociodemographic factors** |  |  |  |
| **Age group** |  |  |  |
|  | 15-24 | 4.6* (4.5, 4.7) | 2.9* (2.9, 3.0) |
|  | 25-44 | 2.8* (2.7, 2.8) | 2.5* (2.4, 2.5) |
|  | 45-64 | 2.1* (2.0, 2.1) | 2.2* (2.1, 2.2) |
|  | 65-84 (Ref.) |  |  |
| **Sex** |  |  |  |
|  | Male | 0.9* (0.9, 0.9) | 1.2* (1.2, 1.3) |
|  | Female (Ref.) |  |  |
| **Income** |  |  |  |
| | \$0-\$39,999 | 2.6* (2.5, 2.6) | 1.2* (1.2, 1.3) |
| | \$40,000-\$59,999 | 1.3* (1.2, 1.3) | 1.1 (1.1, 1.5) |
| | \$60,000 and over (Ref.) | | |
| **Past-year employment** |  |  |  |
|  | Yes | 0.8* (0.8, 0.8) | 0.9* (0.9, 0.9) |
|  | No (Ref.) |  |  |
| **Education** |  |  |  |
|  | Less than secondary school | 1.7* (1.7, 1.7) | 1.5* (1.5, 1.5) |
|  | Secondary school | 1.2* (1.2, 1.2) | 1.3* (1.3, 1.3) |
|  | Post-secondary (Ref.) |  |  |
| **Marital Status** |  |  |  |
|  | Married/Common Law | 0.3* (0.3, 0.3) | 0.5* (0.5, 0.5) |
|  | Single/Widowed/Divorced (Ref.) |  |  |
| **Health factors** |  |  |  |
| **Substance Use** |  |  |  |
|  | Yes | 6.0* (6.0, 6.1) | 3.3* (3.2, 3.3) |
|  | No (Ref.) |  |  |
| **Mood and anxiety disorder** |  |  |  |
|  | Yes | 8.0* (7.9, 8.0) | 2.9* (2.8, 2.9) |
|  | No (Ref.) |  |  |
| **Help-seeking factors** |  |  |  |
| **Seek help** |  |  |  |
|  | Yes | 54.4* (37.0, 80.0) | 32.1* (21.0, 49.0) |
|  | No (Ref.) |  |  |
| *p < 0.05 |  |  |  |

Additionally, those with less education (aOR = 1.5, 95% CI: 1.5, 1.5), lower income (aOR =1.2; 95% CI: 1.2, 1.3) and males (aOR=1.2, 95% CI: 1.2, 1.3) were more likely to be associated with ideation than with no ideation.

There was a weak but statistically significant protective relationship in having a job in the past year and suicidal ideation (aOR = 0.9, 95% CI:0.9, 0.9).

The Hosmer-Lemeshow goodness-of-fit test (p=0.1255) confirmed that the model’s estimates fit the data at an acceptable level.

The second objective sought to differentiate the characteristics of those with ideation alone as compared to those with ideation-with-action. **Table 3.3** provides bivariate comparisons.

**Table 3.3.** Baseline characteristics of those with ideation alone, and those with suicidal plan or attempt

|  | Suicidal ideation alone |  | Suicidal plan or attempt<br>(ideation with action) |  |  |
| --- | --- | --- | --- | --- | --- |
|  | *N*=302 (2.05%) |  | *N*=76 (0.52%) |  | *p* |
|  | *weighted* |  | *weighted* |  |  |
|  | *n* | % | *n* | % |  |
| **Age group** |  |  |  |  | **0.0075<sup>a</sup>** |
| 15-24 | 78 | 26.1 | 28 | 27.8 |  |
| 25-44 | 106 | 40.6 | 19 | 35.9 |  |
| 45-64 | 83 | 25.2 | ** | ** |  |
| 65-84 | 35 | 8.1 | * | * |  |
| 85 and above | 0 |  | 0 |  |  |
| **Sex** |  |  |  |  | **0.6453** |
| Male | 151 | 51.6 | 31 | 41.8 |  |
| Female | 151 | 48.4 | 45 | 58.2 |  |
| **Income** |  |  |  |  | **0.0097** |
| \$0-\$39,999 | 195 | 59.9 | 59 | 72.0 | |
| \$40,000-\$59,999 | 43 | 16.1 | * | * | |
| \$60,000 and over | 29 | 5.7 | * | * | |
| Not stated | 35 | 18.3 | 11 | 21.5 |  |
| **Past-year employment** |  |  |  |  | **0.7023** |
| Employed | 205 | 70.0 | 45 | 74.1 |  |
| Unemployed | 85 | 27.6 | 29 | 25.9 |  |
| **Education** |  |  |  |  | **0.1151** |
| Less than secondary school | 84 | 26.4 | 30 | 31.0 |  |
| Secondary school | 56 | 20.9 | 12 | 9.4 |  |
| Post-secondary | 162 | 52.7 | 34 | 59.6 |  |
| **Marital Status** |  |  |  |  | **0.0002** |
| Married or Common Law | 98 | 42.5 | 15 | 34.5 |  |
| Single/Widowed/Separated/Divorced | 194 | 55.6 | 59 | 64.9 |  |
| **Substance Use** |  |  |  |  | **< 0.0001** |
| yes | 39 | 14.8 | 18 | 18.6 |  |
| no | 256 | 81.8 | 53 | 80.1 |  |
| **Mood or anxiety disorder** |  |  |  |  | **< 0.0001** |
| yes | 87 | 31.9 | 43 | 60.4 |  |
| no | 207 | 67.1 | 29 | 38.6 |  |
| **Substance use & mood or anxiety** |  |  |  |  | **< 0.0001** |
| yes | 14 | 9.7 | 12 | 27.1 |  |
| no | 179 | 90.3 | 23 | 72.9 |  |
| **Seek help** |  |  |  |  | **< 0.0001** |
| yes | 36 | 18.6 | 41 | 71.1 |  |
| no | 266 | 81.4 | 33 | 28.9 |  |
| *Notes:* |  |  |  |  |  |
| *Counts are unweighted.* |  |  |  |  |  |
| *Percentages and p values are weighted.* |  |  |  |  |  |
| <sup>a</sup> *p-value calculated on 15 to 84 age groups.* |  |  |  |  |  |
| * *suppressed due to cell count ≤ 5* |  |  |  |  |  |
| ** *data suppressed so that other small cells cannot be calculated* |  |  |  |  |  |

Overall, 2.1% of respondents (n=302) reported suicidal ideation alone in the past year, and 0.5% (n=76) reporting having ideation-with-action (a plan or a past-year suicide attempt).

The factors that differentiate action from non-action amongst ideators (**Table 3.3**) are remarkably similar to the differences between ideators and non-ideators (**Table 3.1**). Statistically significant bivariate differences were found for all variables except sex, employment and education.

Next, a logistic regression analysis was conducted, examining associations of each covariate with ideation-with-action, adjusting for all the other covariates. Results are shown in **Table 3.4**.

**Table 3.4.**
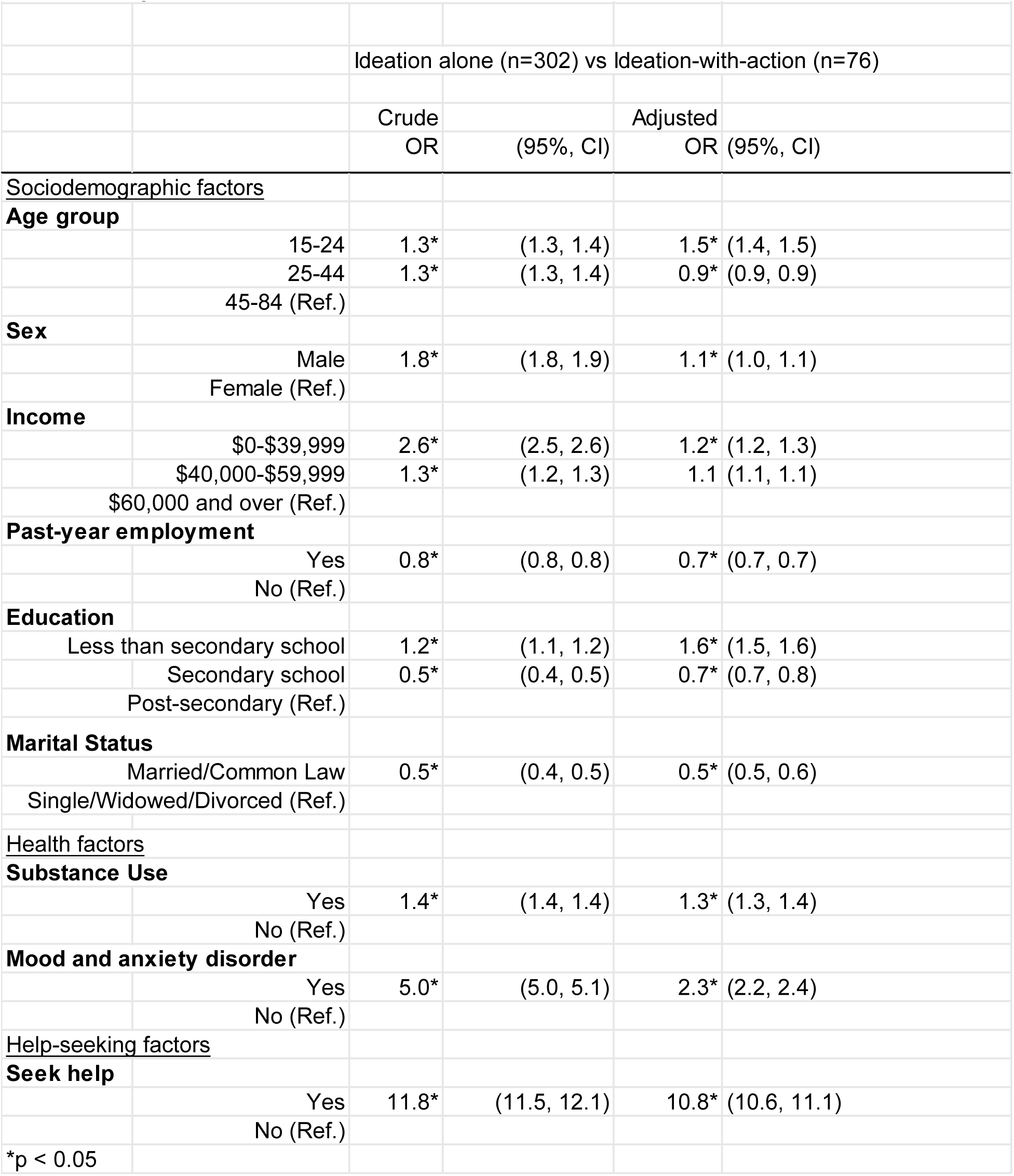
Associations of sociodemographic factors, health conditions, and help-seeking

Even with a relatively small exposure group, all factors were statistically significantly associated with ideation-with-action.

The strongest factors were seeking help (aOR =10.8, 95% CI: 10.6, 11.1), having a mood and anxiety disorder (aOR = 2.3, 95% CI: 2.2, 2.4), less education (aOR = 1.6, 95% CI: 1.5, 1.6), and younger age (aOR = 1.5, 95% CI: 1.4, 1.5). Having a substance use disorder (aOR =1.3, 95% CI: 1.28, 1.4), and lower income (aOR = 1.2, 95% CI: 1.2, 1.3) were also associated with ideation-with-action.

Marriage or common-law relationships were a protective factor against planning/attempt (aOR = 0.5, 95% CI: 0.5, 0.6), as was having a job (aOR = 0.7, 95% CI: 0.7, 0.7).

There was a weak but statistically significant relationships between sex and plan/attempt (aOR = 1.1, 95% CI: 1.05, 1.1).

The Hosmer-Lemeshow goodness-of-fit test (p=0.1) confirmed that the model’s estimates fit the data at an acceptable level.

## Discussion

Suicidal ideation, suicide planning and attempting suicide are a prodrome of suicide deaths (Cao et al, 2015). The study’s finding that 2.6% of community-dwelling respondents aged 15 and above had thought about suicide in the past year equates to about a million Canadians considering ending their lives each year. The rate is in line with the WHO World Mental Health Survey which noted a 12-month prevalence of ideation at 2% and suicide attempts at 0.3% in high-income countries (Borges et al, 2010) including USA (CDC, 2023).

In the Ontario sample, 0.5% also reported having a suicidal plan or a previous suicide attempt in the past year. These proportions are consistent with national proportions in in Canada in 2020: 0.8% of Canadians had planned suicide in the past year, and 0.3% had attempted suicide in the past year (StatsCan, 2023).

### Ideation versus no ideation

When comparing persons with and without ideation, there is agreement with previous studies which found ideators have lower levels of education (Liu et al, 2022; Song and Lee, 2016; Park and Lee, 2015), are poorer (Sueki et al, 2019, Lee et al, 2017, Agerbo, 2007), and are more likely to have substance use disorder and mood and anxiety disorders (Onaemo et al, 2022). Additionally, ideators also tend to be single or divorced or widowed or to have lost connectedness and support (Liu et al, 2022; van Wijingaarden et al, 2014; King and Merchant, 2008; Pompili et al, 2007), consistent with this study’s results. Similarly, the weak but statistically significant protective relationship between having a job and suicidal ideation found in this analysis is consistent with literature (Pirkis et al, 2017; Milner et al, 2014).

Where the results depart from and add to the evidence, is in terms of age and sex: the finding that those with ideation tend to be younger is notable given that there are mixed findings on age as a risk factor in the literature. Some research concurs with this study’s finding that those with ideation tend to be younger (Liu et al, 2022; Cabello et al, 2020) while others found that suicidal ideation has similar prevalence across age groups ranging from 18 to above 65 years old (Rossom et al, 2017).

In terms of sex, the finding that those with ideation tend to be male is consistent with other studies using CCHS data which have found that more than half (56%) of ideators are male (Rhodes and Bethell, 2008) but contrasts with studies that found that those who ideate tend to be female (Zhang et al, 2019; Nowotny et al, 2015) or evenly split between male and female (Liu et al, 2022).

### Ideation alone versus ideation-with-action

When comparing those with ideation alone against those with ideation-with-action (i.e. a plan or previous attempt) there is agreement with previous studies in terms of age: younger age groups are at higher risk of attempting suicide (Kim et al, 2020; Nestman, 2009); marital status: those who are unpartnered are at higher risk (King et al, 2022; Kiecolt-Glaser & Newton, 2001; Dieserud et al, 2001); education: those with lower education have higher risk (Clarke et al, 2014; Dalgard et al, 2007); employment status: the unemployed have higher risk (Skinner et al, 2023; Nordt et al, 2015); and substance use disorder: suicides are ten to 15 times higher in those with substance use disorder compared to the general population (Swann et al, 2022; Ashrafioun et al, 2017; Wilcox et al, 2004).

This study’s finding individuals in the lowest income group had 1.2 times the odds of having action compared to ideation alone, is in line with substantial evidence linking suicide risk and poverty (Drever and Whitehead, 1997; Gunnell et al, 1995). However, there are contrary findings in the literature - a Danish study by Agerbo et al (2001) found that among people with a history of mental illness, those with higher income (rather than lower income) are at greater risk of suicide, and the authors posited that this may be because wealthier people may feel more stigmatised, vulnerable and ashamed about having a mental illness than their counterparts, and this increases their suicide risk.

With regard to mood and anxiety disorders, this study found those who act on their suicidal thoughts were more likely than those with ideation alone to have a mood and anxiety disorder – a finding consistent with studies that have found depression to be the most critical risk factor for attempting suicide (Miller et al, 2020; Yang et al, 2015; Oquendo et al, 2004; Thomas and Morris, 2003). Notably, a cross-national study of 21 countries by Nock et al (2012) found that in developed countries, depression was among the strongest predictors of ideation (OR=3.3) and also predicts the progression from ideation to attempts although the association is less strong (OR=1.5).

The weak relationship between being male and planning/attempt (aOR = 1.03) is consistent with lack of certainty in the literature on sex and suicide risk. A cross-national European study found that suicide attempts were much more likely in males compared to females (Freeman et al, 2017) while others found that suicide attempts are two to four times more frequent among females (Ivey-Stephenson et al, 2022; Bommersbach et al, 2022a; McManus et al, 2016). A particularly notable cross-national study of 85 countries and approximately 85,000 adults found that those who attempt tend to be female (Nock et al, 2008a). Still others have found that sex may not be a statistically significant factor in suicide attempts, as was the case from a nationally representative study in Ireland (Hyland et al, 2021).

With regard to help-seeking, literature generally reports that help-seeking for suicidality is generally low (Calear et al, 2014). This study’s finding adds to this evidence: only 35% of those with ideation (whether acted upon or not) sought professional help for mental health problems.

As expected, among those with ideation, a much higher proportion of those with ideation-with-action sought help (71%) compared to those with ideation-only (19%), which is encouraging and suggests that people with higher severity of ideation and suicidal behaviour are more likely to present at health settings offering care providers an opportunity to intervene.

In general, literature describes suicide ideation as being associated with lower rates of seeking (Ahmedani et al, 2012; Bruffaerts et al, 2011; Deane et al, 2010) and a concept called ‘help negation’ was proffered by Rudd et al (1995) which is a refusal to access help for reasons such as avoiding dependence or perception of threat to autonomy, and poses a major barrier to help-seeking, secondary to stigma from being suicidal (Wilson and Deane, 2010). Some scholars point to low levels of mental health or suicide literacy which is linked to reduced likelihood to seek help (Gabriel and Violato, 2010). Oliffe et al (2016) found that more than half of community-dwelling respondents had poor suicide literacy, particularly men and there were misperceptions in terms of understanding depressive symptoms and suicidality.

Similar to this study’s results, other studies show that help-seeking is relatively more common among those with past suicide attempts – with proportions as high as 92% of those who died by suicide having health care contact prior to death (Schaffer et al, 2016), two thirds of those who attempted suicide contacting their family physician before attempting (Michel et al, 1997, Stenager and Jensen, 1994) and half of those who died by suicide seeking professional help six months prior to death (Chiang et al, 2021).

### Similarities between the two comparison groups

The finding that the comparative risk profiles of the two comparison groups (ideation versus no ideation, and ideation-alone versus ideation-with-action) were the same is in line with a growing body of evidence, meaning that the risk factors that elevate those with no-ideation to ideation are the same that elevate those with ideation only to ideation-with-action. Note, however, that our measurement of the presence or absence of a risk factor does not account for different levels of severity or intensity of a risk factor.

However, the findings concur with May & Klonsky’s (2016) meta-analysis which found that although depression and post-traumatic stress disorder (PTSD) were strongly elevated in ideators compared to non-ideators, and anxiety and drug use disorders moderately elevated in ideators compared to non-ideators, most of these variables did not differentiate attempters from ideators. Depression, alcohol use disorder, hopelessness were similar in both groups and anxiety disorders, post-traumatic stress disorder and drug use disorders were only moderately elevated in attempters compared to ideators. The meta-analysis concluded that there is a continuum of suicidal behaviour and prevention efforts should focus on common risk factors.

Some of this pattern was also found by Kessler et al (1999) in a large epidemiological study which found high odds ratios for distinguishing ideators from non-ideators (e.g. 9.5 for major depression and 14.3 for the presence of three or more clinical disorders) but very low odds ratios for these same factors in distinguishing attempters from ideators (ranging from 1.0 to 2.3), with the authors concluding that all statistically significant factors were more strongly related to ideation, than to progression from ideation to a plan or attempt. The present study found the same patterns in strength of association.

Not seeking help was also a common risk factor in the two comparison groups. (O’Connor et al, 2020). Consistent with Andersen’s Model of Health Care Utilization (Andersen, 1995), enabling and need factors such as stigma and fear of judgment (O’Connor et al, 2020), mental health literacy (Jorm et al, 2010), social support (Thoits, 2011) and perceived need for care (Boudreaux et al, 2010) can facilitate or hinder help-seeking behaviours in people with suicidal ideation as ideation intensifies.

There are a few studies with contrary findings. A study by Nock et al (2010) identified differences between attempters and ideators. Their study showed that having depression increases the odds of being an ideator by 2.3 whereas having depression does not increase the odds that an ideator will attempt suicide (OR = 1.0); additionally, having an anxiety disorder increases the odds of having an attempt by 2.4 whereas it increased the odds of being an ideator by only 1.5.

There could be other reasons for this study’s finding of shared risk factors between the two comparison groups. The study may be missing factors other than the ones measured in the study which were sociodemographic, clinical and help-seeking characteristics. Relevant factors not studied might include genetic predisposition (Poblete et al, 2022; Nielsen et al, 2020) and the nature of suicidal ideation (what and how ideators are thinking, and not only that they did ideate) (Keilp, 2023). Additionally, it was not possible to study crucial factors that drive suicidal behaviour posited in theories. The Interpersonal Theory of Suicide describes the need for acquired capability to make a suicide attempt and individuals being exposed habitually to pain or provocative events such as childhood abuse (Van Orden et al, 2010), and the Integrated Motivational-Volitional Model expands upon this capability concept and describes individuals having access to means prior exposure to suicide, impulsivity and mental imagery (O’Connor, 2011). These factors were not measured in the CCHS surveys. Additionally, there is evidence that distal and proximal factors (Serebriakova et al, 2024; Misiak et al, 2023, Sinyor et al, 2017) may be more important in distinguishing ideators from actors, beyond conventional suicide risk factors, including external events - indeed, self-harm visits were found to have increased during the pandemic in Canada (Gordon et al, 2024).

#### Strengths of the study

Few studies have examined between-group differences in suicidal ideation, as was done in this study. The use of a general population sample generated insight into participants’ sociodemographic characteristics, history of mental health conditions, and help-seeking behaviour and advances understanding in suicidality which incorporates these aspects whereas traditionally, suicide research has tended to focus on clinical/patient or specific populations of interest such as youth, Indigenous communities and veterans.

The use of a large sample allowed for sub-group analyses with sufficient statistical power.

#### Limitations of the study

This study has some limitations. The findings are not directly applicable to subpopulations excluded from the CCHS sampling frame; unrepresented populations include indigenous populations living on reserve, individuals in the military, and those living in institutions. This exclusion is important since it is known, for example, that indigenous persons experience ideation differently (Bolton et al, 2014; Simpson et al, 2003) and indigenous Canadians have three times the suicide rate of non-indigenous Canadians (Kumar and Tjepkema, 2019).

The suicide question in the CCHS were asked only if respondents met a subclinical depression threshold (“Did you feel sad, empty or depressed most of the day, nearly every day, during (a) period of 2 weeks?”). The cohort therefore excludes persons who may have suicidal ideation despite not meeting this threshold. This conditional questioning results in an underestimation of the true prevalence of ideation in the general population.

Persons who did not meet the criteria for the mental disorders covered by the CCHS may still have another type of mental disorder, and others may not have met the criterion of ‘past year’ and could have had disorders earlier in life. Furthermore, the binary categorization used (has or has no mental disorder) does not separate those with severe disorder from those with mild disorder.

The methodology of the study categorizes respondents into ‘non-ideators’, ‘ideators only’ and ‘ideators-with-action’. Treating these categories as fixed may result in underestimating suicide risk in some since suicidality is a phenomenon which is transient and fluctuating (Bornheimer et al, 2022).

The cross-sectional nature of the study design makes it impossible to make causal assertions.

Self-reported mental and substance use disorders are subject to recall bias and have not been verified by clinical diagnosis.

## Conclusion

Persons with suicidal ideation cannot be understood as a homogenous group. They differ in profile in terms of whether they have ideation only, made a suicide plan or have had previous suicide attempts, and in terms of specific sociodemographic characteristics, health status and help-seeking behaviours.

The risk profile of ideators compared to non-ideators is similar to that of ideators-with-action compared to ideators-alone in that more serious groups tend to be male, younger, unpartnered, less educated, have no job, have lower income, have a mood and anxiety disorder, have a substance use disorder and seek help for mental health problems, lending support to the existence of a continuum of suicidality and urging prevention efforts to focus on common risk factors.

Most ideators do not seek help, but planners/attempters are more likely to do so. For those who do present at formal health settings, help must be offered, and for those who do not, suicide literacy, and decreasing stigma and shame may encourage more help-seeking.

## Data Availability

The data are held at Institute of Clinical Evaluative Sciences in Ontario, Canada.

## Appendices

## Appendix 1: Literature justifying the selection of covariates

This study used the following **sociodemographic covariates**: age, sex, marital status, income, education and employment status. Suicide rates have been shown to differ by sex and age: men account for three times the number of suicides than women; suicide rates are highest in adults aged 70 and older across both men and women, and a disproportionately large number take place in younger age groups such as 15 to 29 (WHO, 2014; Patton et al, 2009; Nock et al, 2008a). With regard to ideation specifically (rather than suicide deaths), there is research showing that females had greater levels of suicidal ideation using validated scales, compared to men (Park et al, 2005) but also mixed findings on whether ideation was more prevalent among females or males (Ibrahim et al, 2017; Lewinsohn et al, 2001).

In general, suicide studies do not tend to compare younger to older age groups and instead focus on one age group, highlighting the need for future research since at-risk age groups might differ, especially for cross-country comparisons (Nock et al, 2009). Our study includes logistic regression analyses comparing younger with older age groups and adds to this gap in the literature.

With regard to marital status, literature suggests that separation, divorce and widowhood may increase the risk of suicide (Kolves et al, 2010; Yeh et al, 2008; Brockington, 2001).

With regard to income, research shows that income is inversely associated with psychological distress (McMillan et al, 2010; Lynch et al, 2000; Ferrada-Noli, 1997).

With regard to education, education has been shown to be related to suicidality through the life course in that those with lower levels of education have the highest suicide rates (Phillips and Hempstead, 2017; Lorant et al, 2005).

With regard to employment, a systematic review found that unemployment is associated with greater incidence of suicide (Milner et al, 2014; Milner et al, 2013).

**Mood and anxiety disorders** were investigated as a covariate because of the strength of evidence in the literature on the impact of mood and anxiety disorders on suicidality and its morbid and mortality outcomes. We know that most people with psychiatric conditions do not die by suicide. However, some psychiatric conditions are more strongly associated with suicidal behaviours than others, for example, major depressive episodes account for at least half of suicide deaths (Holma et al, 2014) and is strongly correlated with ideation, attempt and planning (Melhem et al, 2019; Gill et al, 2018; Large, 2016; Nepon et al, 2010; Bertolote and Fleishmann, 2002).

**Substance use and dependence** was investigated as a covariate also because of the strength of evidence in the literature on the impact of substance abuse on suicidality and its morbid and mortality outcomes. Many studies have shown that suicide risk is elevated in connection to substance use disorders, increasing risk of ideation and behaviour. Suicide death is up between ten to 15 times higher in people with alcohol use disorder and opioid use disorder compared to the general population (Swann et al, 2022; Ashrafioun et al, 2017; Wilcox et al, 2004).

Evidence of the relationship between alcohol use and suicide (HHS, 2012; Pompili et al, 2010) exists but is limited. For example, it is known that acute alcohol intoxication is present in about 30 to 40% of suicide attempts (SAMHSA, 2009; Cherpitel et al, 2004). Evidentiary basis for the relationship between drug misuse and suicide is even less explored (SAMHSA, 2015).

There are studies have shown that substance use when present with other mental disorders escalates suicidal behaviour (Nock et al, 2013; Vijayakumar et al, 2011). It is thought that substance use disorder may interact with depression to increase the risk of engaging in suicidal behaviour (Hoertel et al, 2015). Some studies have shown alcohol consumption, specifically, increases the probability of suicide (Edwards et al, 2022; Kim et al, 2021; Rizk et al, 2021) while a meta-analysis found minimal or negligible differences between these two groups (May & Klonsky, 2016) or found mental disorders in general are minimally or negligibly distinguished these two groups (Kessler et al, 1999). Comorbidity of substance use disorder and mood and anxiety disorder as substance use and major depression has been found to increase the risk of suicide attempts (Bachman et al, 2018; Ginley and Bagge, 2017; Beautrais et al, 1996).

Lastly, self-reported **help-seeking** was included a covariate. The question covered help received in the past 12 months for problems with emotions, mental health or use of alcohol or drugs (referred to in the survey as “mental health problems”) and was asked of all respondents. Although it is possible that help-seeking for suicidal ideation may be similar to help-seeking for other mental health issues, there is little research on how help-seeking behaviour might be different for those with suicidal ideation (Ko, 2018). What we do know is that a large proportion of those with suicidal ideation do not seek help. For example, an American study of adults with suicidal thoughts or past suicide attempts found that fewer than two-thirds received treatment in the past year (Ahmedani et al, 2012). This is consistent with a worldwide study that found that fewer than 20% of those with ideation sought help (Bruffaerts et al, 2011).

## Appendix 2: Sensitivity Analyses

Sensitivity analyses were conducted to determine if important differences exist between the 2002 and 2012 surveys, since the study relies on pooling together respondents from these two surveys. Analyses were also conducted to examine the impact of excluding observations with missing data.

### Numbers of respondents self-reporting past-year suicidal ideation

The 2012 survey included respondents who self-reported suicidal ideation during the worst episode of feeling depressed in the universe from which the question of past-year suicidal ideation was asked. The 2002 survey did not. However, the proportion of respondents that had past-year suicidal ideation from 2002 is 24% and this is comparable to the proportion who had past-year suicidal ideation from 2012 (27%) despite the 2002 survey not including the individuals with suicidal ideation during worst/bad episode into the universe for the past-year suicidal ideation question.

### Numbers of respondents reporting seeking professional help

Among those with ideation, in the 2002 survey, the proportion of respondents who reported seeking professional help - which includes visiting ED, hospitalization, walk-in clinic, consulting a family doctor or psychologist in person or by telephone – was 9% (n=20) and in the 2012 survey, that proportion was 29% (n=47). This three-fold increase in proportion who sought help may indicate cultural trends that facilitate help-seeking in the 2012 sub-sample, such as de-stigmatization of mental problems, and is a factor to be mindful of when considering the pooled population of this study.

### Sociodemographic variables

Ensuring the pooled population from the two surveys are not dissimilar in terms of sociodemographic variables is important since we know, for example, that sex is a factor in suicidal behaviour, with higher rates of ideation and attempts among females (Borges et al, 2010; Nock et al, 2008a). In terms of sociodemographic variables, both cycles had similar proportions of respondents in terms of age, sex, marital, and income. In terms of mood and anxiety disorders, and in substance use disorders, 2002 and 2012 survey respondents had similar proportions.

None of the differences above were of concern in terms of affecting the final results of the study.

### Missing data

Categorizing missing sociodemographic variables (age, sex, marital status, income, education and employment) as separate categories for analysis resulted in no meaningful differences.

Where missing data were few, the cases were deleted from the analytic dataset, with no major impact to the result.

## Notes

### Competing Interest Statement

The authors have declared no competing interest.

### Funding Statement

The author(s) received no specific funding for this work.

### Author Declarations

The survey datasets used in this study were accessed at Institute for Clinical Evaluative Sciences (ICES), an independent, non-profit research institute in Ontario which uses population-based health information to produce knowledge on a wide range of healthcare issues. The study cohort consists of Canadian Community Health Survey (CCHS) respondents who already consented to have their survey data shared with ICES for linkage with health administrative data. The use of these data for health system planning and evaluation was authorized under Section 45 of Ontario’s Personal Health Information Protection Act, which allows for administrative approval and does not require review by a research ethics board.

